# ERAP2 protein allotypes show functional diversity in MHC-I antigen presentation in the human population

**DOI:** 10.64898/2026.01.22.26344601

**Authors:** Aroosha Raja, Emma Reeves, Abdulrahman Alasiri, Aafke de Ligt, Alexander Yermanos, Joke H. de Boer, Jeannette Ossewaarde-van Norel, Anastasia Mpakali, Efstratios Stratikos, Marion van Vugt, Edward James, Jessica van Setten, Jonas J.W. Kuiper

## Abstract

The Endoplasmic Reticulum AminoPeptidase 2 (*ERAP2*) gene encodes an aminopeptidase involved in antigenic peptide processing for the MHC-I pathway. Genetic variants in the *ERAP2* gene are associated with autoimmune conditions and infectious diseases. The linkage between genetic variants in the *ERAP2* gene has given rise to the prevailing assumption that a single ERAP2 allotype with invariant amino acid sequence accounts for all immunological functions of ERAP2. Here, we show by analyzing exon-sequencing data from 160,000 individuals that 15 missense amino acid variants across the *ERAP2* gene result in an array of different protein allotypes that are maintained in the European population. These allotypes can be divided into three haplotype groups, defined by the genotypes of two common splice-altering variants. We found novel associations between newly identified protein allotypes and immune-mediated diseases, suggesting that ERAP2 functional variation modifies disease susceptibility at the population level. An MHC class I antigen presentation assay revealed that disease-associated ERAP2 allotypes differ in their capacity to generate antigenic peptides for MHC-I presentation, resulting in differential activation of an antigen-specific T-cell receptor compared to non-disease-associated allotypes. These findings provide strong evidence that ERAP2 function is allotype-dependent and demonstrate that ERAP2 diversity shapes MHC-I antigen presentation and T-cell immunity.

**Significance statement:** The ERAP2 enzyme modulates adaptive immunity and plays a role in autoimmunity, infection, and cancer. The authors discovered that a variety of protein allotypes of ERAP2 are maintained in the human population. Allotypes that increase disease risk for autoimmune and cardiovascular conditions are functionally distinct in their capacity to activate T-cells. The results of this study demonstrate that ERAP2 is a functionally diverse immune modulator that contributes to immune variation and influences susceptibility to immune-mediated diseases.

## Introduction

Endoplasmic Reticulum AminoPeptidases 2 (ERAP2) is an intracellular metalloaminopeptidase involved in the processing of antigenic peptides within the endoplasmic reticulum, facilitating their presentation via the MHC class I pathway (1). By trimming one or more amino acids from the N-terminus of precursor peptides, ERAP2 modifies the sequence composition and binding affinity of peptides to MHC-I and indirectly regulates CD8+ T cell responses (1, 2). Recently, it was shown that ERAP2 also regulates the NKG2A-HLA-E immune checkpoint critical to regulating the cytotoxic activity of natural killer (NK) cells and T cells, by processing the invariant epitopes presented on HLA-E (3). This growing body of evidence suggests that ERAP2 plays a key role in regulating adaptive immunity.

Single nucleotide variants (SNVs) in the *ERAP2* gene influence the splicing and expression of its encoded enzyme, thereby affecting its role in the regulation of adaptive immunity (1). Recent genetic studies of ancient populations suggest that these variations in the *ERAP2* gene may have been subject to positive selection due to their role in enhancing resistance to lethal infectious diseases (1, 4–6). These same variants are also associated with increased protection against contemporary infections, including HIV, COVID-19, and bacterial pneumonia (7, 8). Intriguingly, the SNVs linked to protection against infectious diseases are also associated with a heightened risk of developing severe autoimmune conditions (1, 8–10). Furthermore, these SNVs have been linked to altered outcomes of cancer immunotherapy (3), pregnancy complications (11), and blood pressure (12). Despite these associations, the mechanisms by which *ERAP2* gene variations influence susceptibility to these diverse pathologies remain unclear.

There are hundreds of SNVs in the *ERAP2* gene, but due to genetic linkage (also known as linkage disequilibrium, LD, i.e., SNVs inherited together), the current understanding is these SNVs assemble in two major haplotypes, determined by the genotype of a single SNV, rs2248374. The haplotype containing the A allele of rs2248374 encodes *full-length* protein (∼100 kDa), whereas the alternative haplotype with the G allele disrupts a canonical splice site and introduces a premature stop codon. This results in truncated mRNA that is degraded by nonsense-mediated mRNA decay (NMD) and does not produce protein (1, 10, 13). The distribution of these two haplotypes (one produces protein, the other does not) in the population is thought to account for the disease predisposition associated with ERAP2. Consequently, all biological functions of ERAP2 are attributed to a single protein isoform (1, 4, 5, 8).

However, we and others have discovered that the association of variants in *ERAP2* with autoimmune diseases involve uncharacterized haplotypes independent of rs2248374 (10, 14). Furthermore, it has been reported that, under certain inflammatory conditions, the ‘loss-of-function’ haplotype (with the G allele of rs2248374) may produce a short ERAP2 protein of about half the size (∼50kDa) (15–17). These findings suggest that the widely believed dichotomous (“all or nothing”) haplotype model of *ERAP2* may not accurately describe the functional diversity of ERAP2 haplotypes in the human population. To date, genetic association studies have linked ERAP2 haplotypes with diseases tested for individual single nucleotide variants (SNVs), resolving haplotypes based on LD with known functional SNVs. This approach may overlook haplotypes formed by combinations of SNVs with varying allele frequencies in the population (18, 19).

Note that missense SNVs in the homologous and functionally complementary gene *ERAP1* assemble in multiple haplotypes in the human population that exhibit a wide range of enzymatic activities towards peptide substrates and differentially shape the immunopeptidome of MHC-I (9, 20–23). These protein-altering haplotypes are referred to as “allotypes” because they change the enzyme’s function and may therefore alter immune responses; notably, different ERAP1 allotypes can be protective or confer risk depending on the specific disease, resulting in disease-specific and sometimes opposing associations (9, 20, 23, 24). A recent study testing SNVs that change ERAP2 amino acid sequences in the 1000 Genomes Project, demonstrated that similar to ERAP1, different ERAP2 allotypes persist with altered frequencies across continental populations (1). However, a detailed analysis of the function of ERAP2 allotypes and their association with human disease traits is currently lacking. Information on naturally occurring ERAP2 protein allotypes may more accurately recapitulate the complexity of *ERAP2* function in the contemporary human population, and potentially reveal allotype-specific disease associations (9, 25, 26). Deconvolution of the biological properties of ERAP2 into distinct protein allotypes, may open the door to intelligently designed approaches targeting specific allotypes to modulate ERAP2 activity in diseases without compromising normal functions. In this study, we analysed exome sequencing data from 160,000 individuals of the UK Biobank to characterise the ERAP2 allotypes in a human reference population and determined their association with several human disease traits and compared their aminopeptidase function using a cell-based antigen presentation assay.

## Methods

### Exome-sequence data from the UK Biobank

Exome-sequencing data from the UK Biobank was obtained under application no. 24711. Details on the exome sequencing in DNA samples from the UK Biobank study have been described elsewhere (27). Briefly, the exomes were captured using 5’ biotinylated, oligonucleotide probes (IDT xGen Exome Research Panel v1.0) and sequenced with dual-indexed 75 x 75 bp paired-end reads on the Illumina NovaSeq 6000 platform using S2 and S4 flow cells. See for further details the UK Biobank documentation at https://www.ukbiobank.ac.uk/media/najcnoaz/access_064-uk-biobank-exome-release-faq_v11-1_final-002.pdf.

### ERAP2 haplotype estimation

Whole-exome sequencing data (ancestry-specific files in *Plink2* PGEN format) from 160,000 individuals of European descent (previously assigned based on array data released by the UK Biobank study to determine continental ancestry super-groups) in the UK Biobank were subjected to de novo phasing without a reference genome using *Eagle2* for common variant (minor allele frequency [MAF] >0.1%) phasing. To accurately phase rare variants (MAF<0.1%), *Eagle2* uses haplotype scaffolds derived from common variants. Phased exome-sequencing data were filtered for genotype data in chromosome *5q15* spanning the *ERAP2* locus (build GRCh38, chromosome 5 from position 96875986-96919703, forward strand) using *BCFtools* in *Python3*. We identified rsID’s of SNVs by cross-referencing positions of SNVs with data from the gnomAD v3.1.2 database (28). Genotype binary dosages were converted into reference and alternative nucleotide alleles for each variant and the frequency of each variant was calculated by *Plink 2.0* (29). SNVs were filtered against missense variants reported by gnomAD v3.1.2 and an allele frequency threshold ≥ 0.0001 [1:10,000].

Predictions for all possible single amino acid substitutions in ERAP2 by AlphaMissense (30) were obtained by filtering for Uniprot accession Q6P179 in the *“AlphaMissense_aa_substitutions.tsv.gz”* file at the AlphaMissense repository at https://zenodo.org/records/8208688.

### *ERAP2* splice variant selection

There are two common SNVs in *ERAP2*, s2248374 and rs17486481, that can cause alternative splicing of mRNA (1). To identify additional splice altering SNVs in high linkage disequilibrium (LD) with missense variants used for *ERAP2* haplotype estimation, we first calculated the LD [r^2^] with the --*LD* function in *Plink 2.0* between the missense variants identified in this study (n=15) and variants from the whole genome sequencing data set of the UK biobank. We also obtained nucleotide positions predicted to affect splicing in the *ERAP2* gene using *SpliceAI* (31) using precomputed scores (“spliceai_scores.raw.snv.hg38.vcf.gz” available via https://basespace.illumina.com/analyses/194103939/files?projectId=66029966 and the linux command (MacOS): *zgrep ‘ERAP2’ spliceai_scores. raw. snv. hg38. vcf. gz > spliceai_scores. raw. snv. hg38. vcf*.) to filter for variants in *ERAP2.* We then cross-referenced *ERAP2* SNVs with high LD(r^2^>0.7) with the missense variants identified by WES with SNVs with spliceAI scores. The expected frequency of haplotypes under varying levels of LD (r^2^) and allele frequencies (assuming biallelic loci with symmetric allele frequencies or simplicity) was modelled in R version 4.4.0. The ancestral haplotype of splice variant rs2248374 and missense variant rs2549782 was calculated using the allele frequencies and LD (r^2^) of these variants in the whole genome sequencing data set of the UK biobank using the formula:

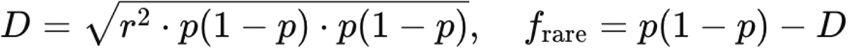

### Multiple sequence alignment and phylogenetic analysis

Multiple sequence alignment of *ERAP2* haplotypes was done using CLUSTAL W (32) with default parameters using Biopython libraries in Python 3. For evolutionary analysis, the *Neighbour–Joining* (NJ) method was used to construct phylogenetic trees (33). The tree was constructed using following steps:

1. Calculating distance between sequences to formulate a distance matrix
2. Calculating distance between OTU one with all other OTUs

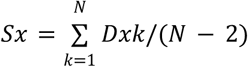 #N=operational taxonomic units (OTUs) and S is the sum of the distance (D)
3. Identifying the OTUs pair with smallest distance

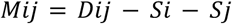
4. Calculating branch length

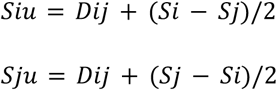
5. Round2 til Round x, calculating a new distance matrix by connecting i and j according to S (4) and replacing it with a node (u). Calculating new distance matrix of all other tax to u with

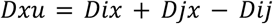

The tree was rooted to the ancestral allotype recently termed *ERAP2*00:01* (1). The tree was constructed using *scikit-bio* (from: scikit-bio.org) and *ete3* (34) libraries in python3.

### Disease association analysis in the UK Biobank

For disease association we used the 384 phenotypes of the ICD-10 (International Classification of Disease, 10th Revision) codes in the UK Biobank. The full list of the phenotypes investigated were obtained from UK Biobank under application no. 24711. The association of *ERAP2* haplotypes with the phenotypes was assessed by logistic regression model using *statsmodels* library in *Python* (35). Broyden–Fletcher–Goldfarb–Shanno (BFGS) optimization method was used to avoid any convergence issue in the logistic regression model. Nominal p-values were corrected for type-1 error using the Benjamini–Hochberg (BH) method. The calculated odds-ratio, corresponding 95% confidence interval, adjusted *p* values are reported in the **Supplementary Table 1**.

### Site-directed mutagenesis

The mutagenesis reactions to introduce the L669Q substitution into ERAP2 was performed using the QuickChange II Site-Directed Mutagenesis Kit (Agilent Technologies), according to the manufacturer’s instructions. The sequences of primers used were: L669Q (t2006a); forward primer; 5’-tcatgatgtgtttcagcaagttggtgcagggagac-3’, and L669Q (t2006a); reverse primer; 5’-gtctccctgcaccaacttgctgaaacacatcatga-3’. After the mutagenesis reaction, the selected mutation was verified by sequencing (VBC Genomics, Austria).

### Protein expression and purification

ERAP2 protein and variants were expressed using the Bac-to-Bac expression system (Invitrogen Life Technologies) according to the manufacturer’s instructions as described before (36). Recombinant ERAP2 enzymes were expressed by infecting Hi5 insect cell cultures (50–100 mL) with the P2 baculovirus stock at a volume ratio of 1:25 to 1:50 (virus to culture). Culture supernatants were harvested 72 hours post-infection. The clarified supernatant was incubated with Ni-NTA resin in phosphate buffer (pH 7.0) supplemented with 20 mM imidazole to minimize non-specific binding. After washing, bound protein was eluted using a stepwise imidazole gradient. Elution fractions (1–1.5 mL) were collected and screened for enzymatic activity using a kinetic assay that monitors the hydrolysis of a synthetic fluorogenic peptide substrate. Active fractions were dialyzed overnight against 10 mM HEPES, 100 mM NaCl, pH 7.0. Final protein concentrations were determined by densitometric analysis of SDS-PAGE gels, using a reference protein of known concentration as a standard.

### *In vitro* enzymatic assays

The aminopeptidase activity of recombinant ERAP2 was assessed using a fluorogenic substrate L-Arginine-7-amido-4-methylcoumarin (R-AMC; Sigma-Aldrich). Hydrolysis of L-Arginine-7-amido-4-methylcoumarin was monitored by measuring fluorescence change over 15 minutes (excitation at 380 nm, emission at 460 nm). Reactions were carried out in a buffer containing 20 mM HEPES (pH 7.0), 150 mM NaCl, and 0.002% Tween-20. Fluorescence measurements were performed using a TECAN Infinite M200 microplate reader. Specific activity was calculated as mol of product (AMC) formed per mol of enzyme per second, using a standard curve generated with known concentrations of AMC. Reaction rates were determined from the slope of the fluorescence or absorbance signal over time. For Michaelis-Menten analysis, reaction rates were measured across a range of substrate concentrations up to 150 μΜ. Substrate dilutions were prepared at threefold the final concentration in Eppendorf tubes, and 50 μL of each dilution was transferred in duplicate to a 96-well microplate. The enzyme was diluted in reaction buffer (20 mM HEPES, pH 7.0, 150 mM NaCl, 0.002% Tween-20) to achieve a final concentration of 10 nM upon addition of 100 μL to each well. Initial reaction rates were determined from the linear portion of the kinetic curves. Specific activity values (reaction velocity per enzyme concentration) were calculated for each substrate concentration, and the resulting data were analyzed using GraphPad Prism 8.0. Fits were applied using the classical Michaelis-Menten model.

### T-cell activation assay

ERAP1 and ERAP2 genes were knocked out using CRISPR-Cas9 in HEK293T cells (*293T-ERAP-KO*). Cells were authenticated by STR analysis and were confirmed Mycoplasma free. For assessment of extended precursor SIINFEHL (R-SHL8) trimming, 293T-ERAP-KO cells were transfected using FuGENE 6 Transfection Reagent (Promega) and 1μg total DNA, consisting of DNA constructs encoding non-functional ERAP2 (E337A) or naturally-occurring *ERAP2* allotypes (0.25μg), alongside H2-K^b^ (0.25μg) and the N-terminally extended ERAP2 specific R-SHL8 (0.5μg), as described before (37). The transfected cells were incubated for 24 hours at 37°C/5% CO_2_ before harvesting and overnight co-culture with the H2-K^b^ restricted SHL8-specific, LacZ inducible T-cell hybridoma, B3Z (37). Presentation of final SHL8 peptide and activation of the LacZ inducible T cell hybridoma was assessed by measurement with the substrate chlorophenolred-b-D-galactopyranoside (CPRG) by its absorbance at 595 nm and 655 nm as reference. Trimming of R-SHL8 by ERAP2 and ERAP2 variants was indirectly measured through B3Z activation and the induced CPRG colour change.

### Data availability

All exome-sequencing data used in this research are publicly available to registered researchers through the UK Biobank data-access protocol under application no. 24711. The resources including software, reagents, summary statistics and scripts used to analyse the data are available in the **Supplementary Table 2** and github repository https://github.com/Araja1234/ERAP2-Haplotypes-Analysis-UK-Biobank/

## Results

### Protein-altering variants in the *ERAP2* gene

To accurately characterise *bona fide* ERAP2 allotypes, we used a conservative strategy. We first analysed whole-exome sequencing (WES) data from 160,000 individuals of the UK Biobank (UKB). We performed de novo haplotype phasing of WES data using the common variants (Minor allele frequency [MAF] above 0.1) and phased rare variants onto the resulting haplotypes by imputation. Next, we limited our analysis to known missense variants in the *ERAP2* gene with sufficient coverage. To achieve this, we first filtered for missense variants reported in the *gnomAD* database with a minor allele frequency (MAF) ≥0.0001 (1:10,000) in the WES data, so each variant occurred at least 32 times (**Figure 1A, Supplementary Table 3**). Following filtering, 15 missense variants were identified in exon 2 through exon 18 of the *ERAP2* gene (**Figure 1B**), with allele frequencies closely matching reference data for the European population in gnomAD (**Figure 1C**). Based on the 3-dimensional structure of the ERAP2 protein, the 15 missense variants were positioned throughout the ERAP2 protein, including near the catalytic site(**Figure 1D**).

**Figure 1.**
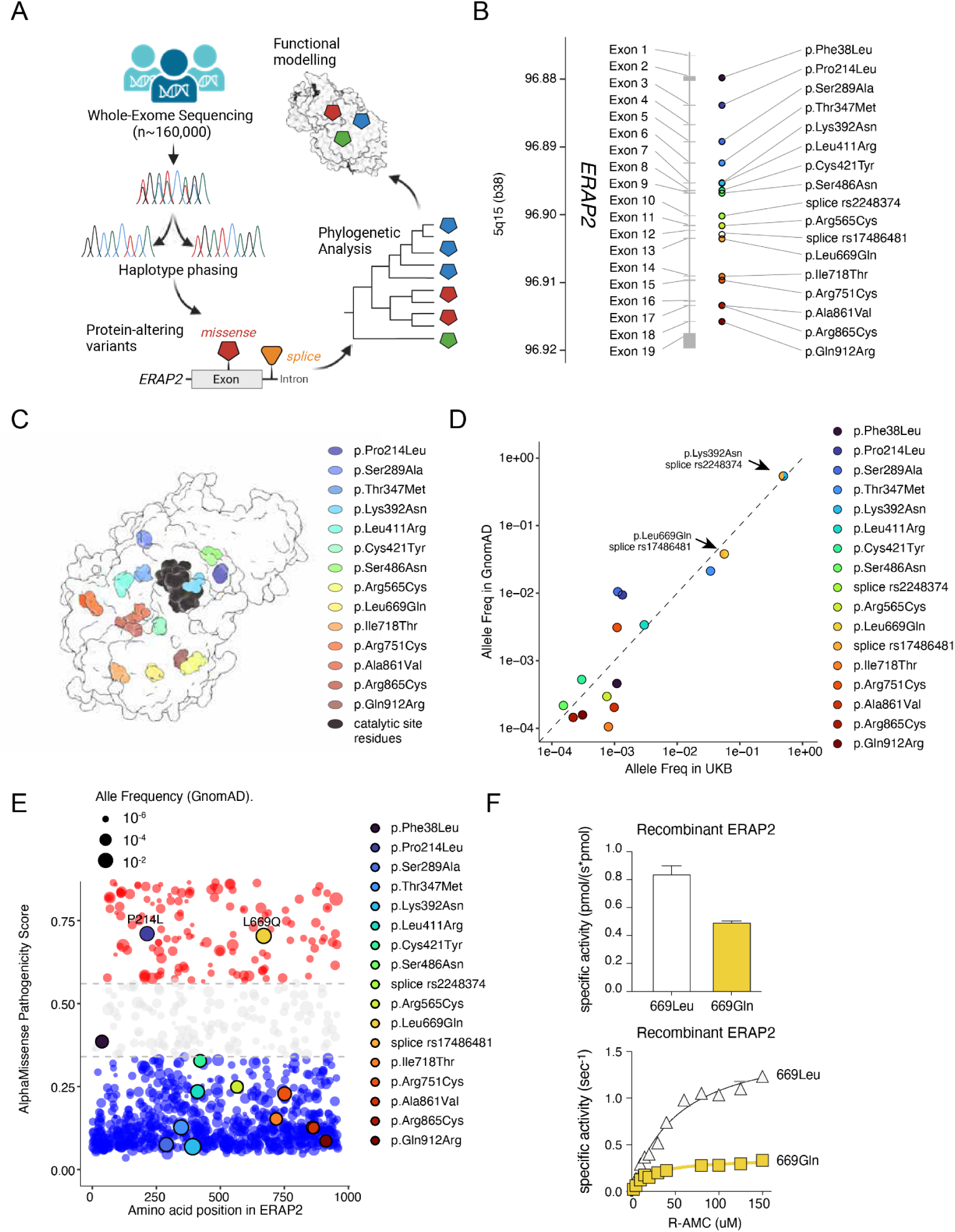
The *ERAP2* gene maintains seventeen protein-altering variants in the European population. **A**) Study design: In this study, we identified ERAP2 haplotypes using single nucleotide variants (SNV’s) from exome-sequencing data from the UK Biobank (UKB). We compared haplotypes phylogenetically, evaluated their association with disease phenotypes in the population, and evaluated their functional differences. **B**) We identified 15 missense variants and two splice-altering SNVs at an allele frequency ≥0.0001 in the UKB that map across the *ERAP2* gene body. **C)** The ERAP2 protein structure (PDB ID: 7SH0 in ChimeraX) highlighting the amino acid positions encoded by the 15 missense variants in *b*, as well as the zinc binding catalytic site residues (His370, His374, and Glu393) that form the HEXXH(X)18E zinc-binding motif of ERAP2. **D**) The allele frequency of the 17 SNVs in the UKB in this study and in the Genome Aggregation Database (gnomAD). The allele frequency of the intronic splice-altering variant rs17486481 was estimated from p.Leu669Gln (LD [r^2^]∼1.0). **E)** AlphaMissense pathogenicity scores (0–1) of predictions for all possible single amino acid substitutions for 3965 annotated variants in ERAP2 (gnomAD version 4, build 38). The AlphaMissense scores represent the predicted probability of a variant being clinically pathogenic. Variants are classified by AlphaMissense as *likely benign* (<0.34, in blue), *likely pathogenic* (>0.56, in red), or *ambiguous* (in between, in grey). The 15 missense variants identified in *b* are highlighted. A substitution’s size is based on the allele frequency of the variant at that position in gnomAD. **F**). Specific activity of recombinant ERAP2 with the 669L versus 669Q variant in the trimming of the fluorogenic substrate Arginine-7-amido-4-methylcoumarin (R-AMC). Panel B, Michaelis-Menten analysis of ERAP2 with the L669 versus 669Q variant for the trimming of R-AMC.

Protein expression of *ERAP2* is strongly regulated by two common SNVs causing alternative splicing (1). The WES data we used for haplotype phasing may miss additional intronic splice altering variants in strong LD with the 15 missense variants. To find out if this was the case, we calculated the LD between the 15 missense variants and all *ERAP2* variants with MAF ≥0.0001 in *whole-genome-sequencing* data from the UKbiobank that became available during the course of this study. After filtering variants in LD [r^2^>0.7, n=123] with the 15 missense variants against loss-of-function annotations (pLOF) in gnomAD (i.e., stop gained, splice altering, frame shift. etc) or scores from the splicing prediction algorithm *spliceAI* (thresholds for splicing was set at a liberal >0.3), two of the missense variants were in near full LD with the known splice variants: splice variant rs2248374 with rs2549782 (r^2^ = 0.999391) and splice variant rs17486481 with rs17408150 (r^2^ = 0.999591). These splice-altering variants define Haplotypes A-C of *ERAP2* (1). We did not identify any other splice-altering variants in LD with the missense variants. As a result, haplotypes encoding the A allele of rs17408150 (669Q) were considered to have the G allele of rs17486481. Data for rs2248374 were present in the exome-sequencing data because its position is immediately downstream of exon 10, and thus we included the genotype for this variant in our further analyses.

The G allele of the deep intronic variant rs17486481 introduces a 5’ splice site and exposes a premature stop codon [SpliceAI, donor gain score = 0.53]) (1, 38). The 669Q (rs17408150) allele shows a strong negative association with ERAP2 plasma levels (GWAS catalogue accession: GCST90241043, rs17408150-A; *P* = 4 x 10^-230^) which suggests that haplotypes encoding the splice-altering variant rs17486481 may reduce ERAP2 protein expression. Since the G allele of splice-altering variant rs2248374 is in strong LD with the T allele of rs2549782 (392N), which prevents the expression of an ERAP2 variant with substantially altered ERAP2 enzymatic activity (39, 40), we hypothesized that the strong LD between the G allele of rs17486481 and 669Q also suggests that 669Q significantly affects the enzymatic activity of ERAP2. To assess this, we first used the deep-learning model *Alpha2Missense* that combines structure prediction with AlphaFold2 and protein language modelling against allele frequencies in the human population to provide state-of-the-art variant effect predictions. Based on Alpha2Missense results, the L669Q variant is “likely pathogenic”, indicating that it is a disruptive amino acid substitution (**Figure 1E**). To confirm this experimentally, we generated a recombinant ERAP2 protein variant carrying the 669Q substitution and compared its enzymatic activity to the wild-type ERAP2, which contains a Leucine (L669) at that position. The L669Q substitution resulted in a 60% reduction in specific enzymatic activity (**Figure 1F**), supporting that this variation significantly alters the activity of ERAP2. The Michaelis-Menten analysis revealed that although 669Q results in a slight increase in substrate affinity (the kinetic parameter K_M_), the decrease in specific activity is mostly due to a marked decrease in catalytic efficiency (as indicated by the k_cat_ parameter) (**Supplementary Figure 1**).

In summary, an analysis of exome sequencing data in ∼160,000 individuals identified 15 missense variants and two splice-altering variants in the *ERAP2* gene in the European population.

### There are 10 protein-coding ERAP2 allotypes in the European population

Next, we considered the phased haplotypes of the 15 SNVs and the 2 essential splice variants which identified 27 *ERAP2* haplotypes in the European population (**Figure 2A-B**). We classified each of these haplotypes according to standardised nomenclature previously proposed for *ERAP2* haplotypes based on data from the 1000 Genomes Project (1). Phylogenetic tree analysis of the 27 haplotypes using the ancestral haplotype as the root node (**Figure 2B**) revealed three large distinct clades that represent the 3 major haplotypes (Haplotypes A-C) controlled by the splice variants (**Figure 2A**). The G allele of rs2248374 tags 13/27(48%) haplotypes while rs17486481 tags *ERAP-2*10:01-10:03, and ERAP-2*11:02* (4/27, 15%). Consequently, 10 ERAP2 haplotypes were considered capable of producing full-length ERAP2 (10/27, 37%), which we will refer to as ‘allotypes’.

**Figure 2.**
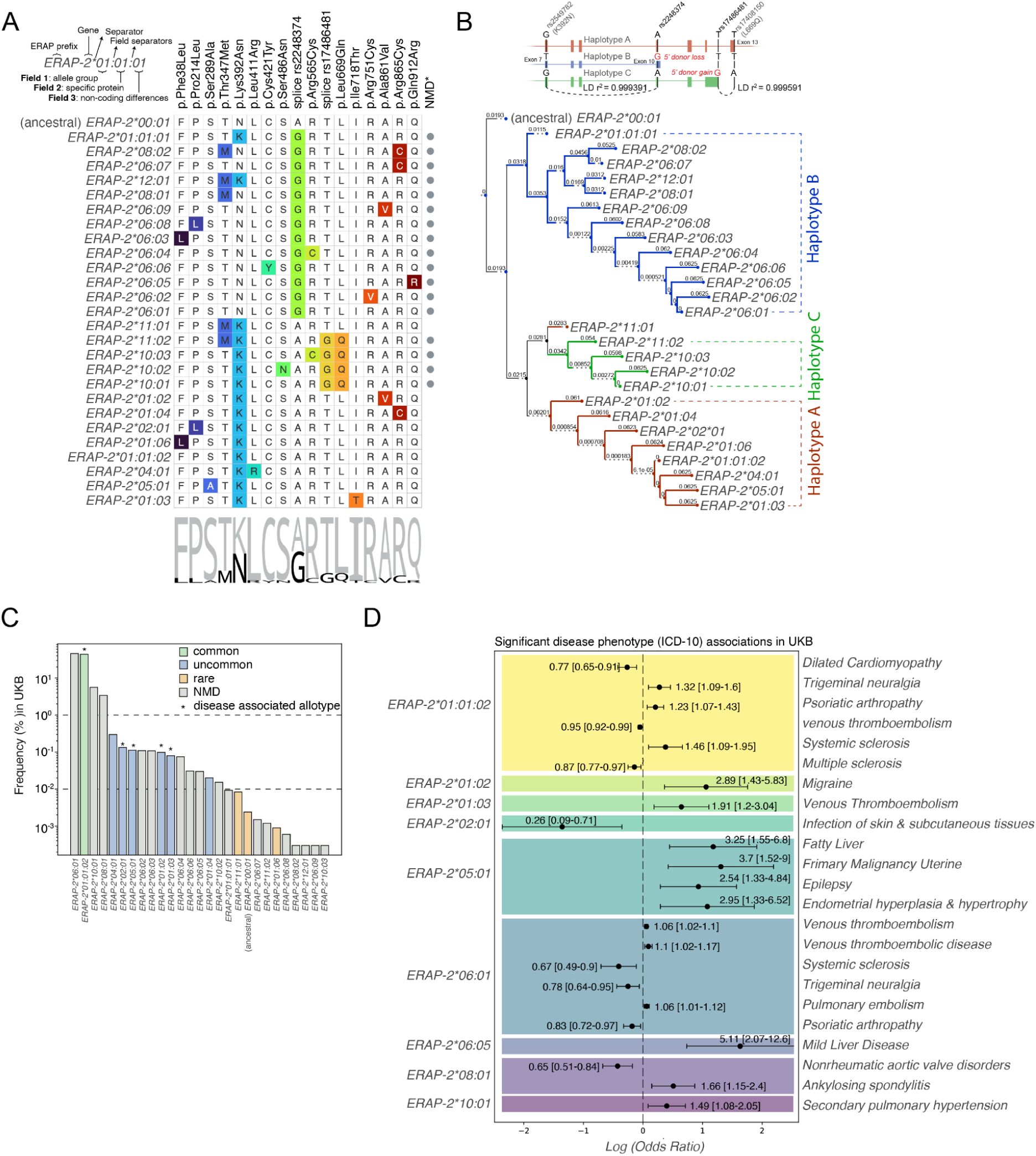
Population-level diversity in ERAP2 protein allotypes modulates susceptibility to various disease phenotypes. **A)** Multiple sequence alignment of severenteen phased missense and splice-altering SNVs distinguishes 27 ERAP2 haplotypes. The haplotypes were classified according to standardised nomenclature (1). The amino acid substitutions (except for the splice-altering variants) encoded by the alternative allele are highlighted and summarized by a sequence logo. Haplotypes predicted to undergo nonsense mediated mRNA decay (NMD) are highlighted by grey dots. **B**) Rooted cladogram constructed using the Neighbor-joining (NJ) method of the 27 haplotypes using the ancestral haplotype as the root node, revealed three clades that represent the 3 major haplotypes (Haplotypes A-C) controlled by the two splice variants rs2248374 and rs17486481. The branch distances (lengths) in the tree are shown and were calculated step-by-step using the NJ method, which builds the tree by repeatedly joining the closest nodes based on their genetic distance. The haplotype model for ERAP2 with the splice variants and missense variants in strong LD (r^2^) are indicated at the top. **C)** The frequency (%) of the 27 haplotypes in the UKB WES data (n ∼160,000) identified in this study. Haplotypes frequency was classified as common (>1%), uncommon (1-0.01%), or rare (<0.01%). Haplotypes predicted to be susceptible to nonsense mediated mRNA decay (NMD) as shown in *b* are highlighted in grey. Protein allotypes (i.e., full-length encoding ERAP2 protein haplotypes) that show significant association with ICD-10 disease phenotypes in *d* are indicated by asterisks. **D)** Disease association of ERAP2 haplotypes in UK Biobank data. Associations were tested across ICD-10 (the International Classification of Diseases, Tenth Revision) phenotypes, and significant results (Bonferroni-corrected p-values) are shown. The x-axis is scaled using the logarithm of the odds ratio (OR), and the calculated ORs with 95% confidence intervals are displayed next to each significant ICD-10 diagnostic code.

We noted that 4 haplotypes were relatively common (>1%) of which one is the “major” allotype *ERAP-2*01:01:02* (**Figure 2C**). We detected 12 haplotypes that were uncommon (1-0.01%) of which 6 were allotypes, including *ERAP-2*02:01*, *ERAP-2*04:01* and *ERAP-2*05:01,* and 11 were rare (<0.01%) in the European population (**Figure 2C**). In agreement with our recent analysis of the 1000 Genome Project data (1), the African alleles *ERAP-2*03*, *ERAP-2*07*, and *ERAP-2*09* were absent from the European population in the UK Biobank. Interestingly, the ancestral allotype characterised by asparagine at amino acid position 392 [392N], which alters ERAP2 substrate specificity (39), was previously thought to be exclusive to contemporary non-human primates (1, 13). Yet, our data suggests it may be maintained in the European population at low frequency (haplotype frequency of 2.395 × 10^-5^ or ∼0.0024% of ERAP2 haplotypes, **Supplementary Table 3**) (**Figure 2C**). This is very close to the estimated frequency of 8.9 ×10^−5^ for this haplotype based on modelling the allele frequencies and LD between variants rs2248374 and rs2549782 in the UK Biobank (**Supplementary Figure 2**).

### ERAP2 haplotypes and human traits

Next, we asked if the identified ERAP2 haplotypes are associated with human traits in the UK Biobank (UKB). To this end, we conducted haplotype-based association analysis using 382 ICD-10 phenotypes of the UKB which were further filtered for duplicates and N/A clinical outcome names (179 final phenotypes), identified several significant disease associations for ERAP2 allotypes (BH adjusted *P* value<0.05). As expected, most associations for were detected for the common allotype *ERAP2*01:01:02*, which ascertained the increased risk for several autoimmune diseases, such Psoriatic arthropathy, more commonly known as Psoriatic Arthritis (PsA)(odds ratio (OR) [95% CI]=1.23[1.07-1.43], *Padj* = 1.41 × 10^-2^) and Systemic sclerosis (OR [95%CI]: =1.46[1.09-1.95], Padj = 3.42 × 10^-2^), in line with previous observations for these conditions (5, 41). We further identified novel disease associations, such as between *ERAP2*01:01:02* and Trigeminal neuralgia and protection against venous thromboembolism (**Figure 2D, S2 Supplementary Table 1**). In contrast, the *ERAP2*01:03* allotype (characterised by threonine at amino acid position 718 [718T]) was significantly associated with increased risk for Venous thromboembolism (OR [95%CI]: =1.91[1.2-3.04], Padj = 2.02 × 10^-2^)(**Figure 2D**). Furthermore, the allotype *ERAP2*01:02* (characterised by valine at amino acid position 861 [861V]) was significantly associated with increased risk for migraine (OR [95%CI]: =2.89[1.43-5.83], Padj = 9.42 × 10^-3^), and *ERAP2*02:01* (characterised by leucine at amino acid position 214 [214L]) with protection against Infection of skin and subcutaneous tissues (OR [95%CI]: =0.26[0.09-0.71], Padj = 2.55 × 10^-2^). Finally, the allotype *ERAP2*05:01* (characterised by alanine at amino acid position 289) was significantly associated with increased risk for epilepsy, liver disease and uterine or endometrial pathology (**Figure 2D**). We noted that several allotypes are associated with altered risk for venous thromboembolism. This finding is interesting since a large GWAS meta-analysis associated a very strong ERAP2 eQTL (rs1160962, GTEx Whole blood *P* = 1.8 × 10^-267^) with increased risk for venous thromboembolism (42). We conclude that specific ERAP2 allotypes are associated with altered susceptibility to a variety of human diseases, in particular immune-mediated and inflammatory conditions, a finding that is consistent with the established biological role of ERAP2.

### Disease-associated ERAP2 allotypes are functionally distinct

To assess the trimming function of the disease-associated ERAP2 allotypes identified, we used the well characterized H-2K^b^-SIINFEHL (SHL8) antigen presentation model system. To this end, ERAP-deficient HEK293T (by CRISPR-Cas9 mediated knock-out of *ERAP1* and *ERAP2* genes) were transfected with a DNA constructs encoding four disease-associated or four control ERAP2 allotypes along with N-terminally extended precursor R-SIINFEHL (R-SHL8) with an ER translocation signal. The expression of trimmed SHL8 presented by H2-K^b^ (MHC-I) at the cell surface was measured by co-culturing transfected cells with the H2-K^b^/SHL8-specific T cell hybridoma B3Z at different effector:target ratios, allowing direct assessment of the trimming activity of ERAP2 allotypes via the activation of an antigen-specific T cell receptor (**Figure 3A**). Transfection of the major *ERAP2*01:01:02* allotype resulted in a strong dose response and was used as a reference for ERAP2 trimming activity (**Figure 3B**). A negative control was created by mutating the E337 amino acid in the GAMEN motif to 337A to produce an ERAP2 allotype without aminopeptidase activity (ERAP2-GAMAN; **Figure 3B**). A comparison of the trimming capacity of the selected four disease-associated ERAP2 allotypes (ERAP2*01:02, ERAP2*01:03, ERAP2*02:01, and ERAP2*05:01) and the four non-disease-associated allotypes (the ancestral ERAP2*00:01, ERAP2*01:06 [38L], ERAP2*04:01 [411R], and ERAP2*11:01 [347M]) revealed that disease-associated allotypes were more efficient in trimming R-SHL8 compared to the major allotype (*P*<0.05). In contrast, control allotypes showed a capacity relatively similar to the major allotype (*P*>0.05, **Figure 3C**). The ERAP2*05:01 (289A) (*P* = 1.14 × 10⁻³) and ERAP2*01:02 (861V) (*P* = 9.93 × 10⁻³) variants exhibited the highest ability to generate the mature epitope, with both showing ∼145% of the major allotype activity (**Figure 3B and C**). Overall, our findings indicate that the discrete ERAP2 allotypes linked to altered disease susceptibility in the human population exhibit significantly altered enzymatic activity compared to the major allotype of ERAP2.

**Figure 3.**
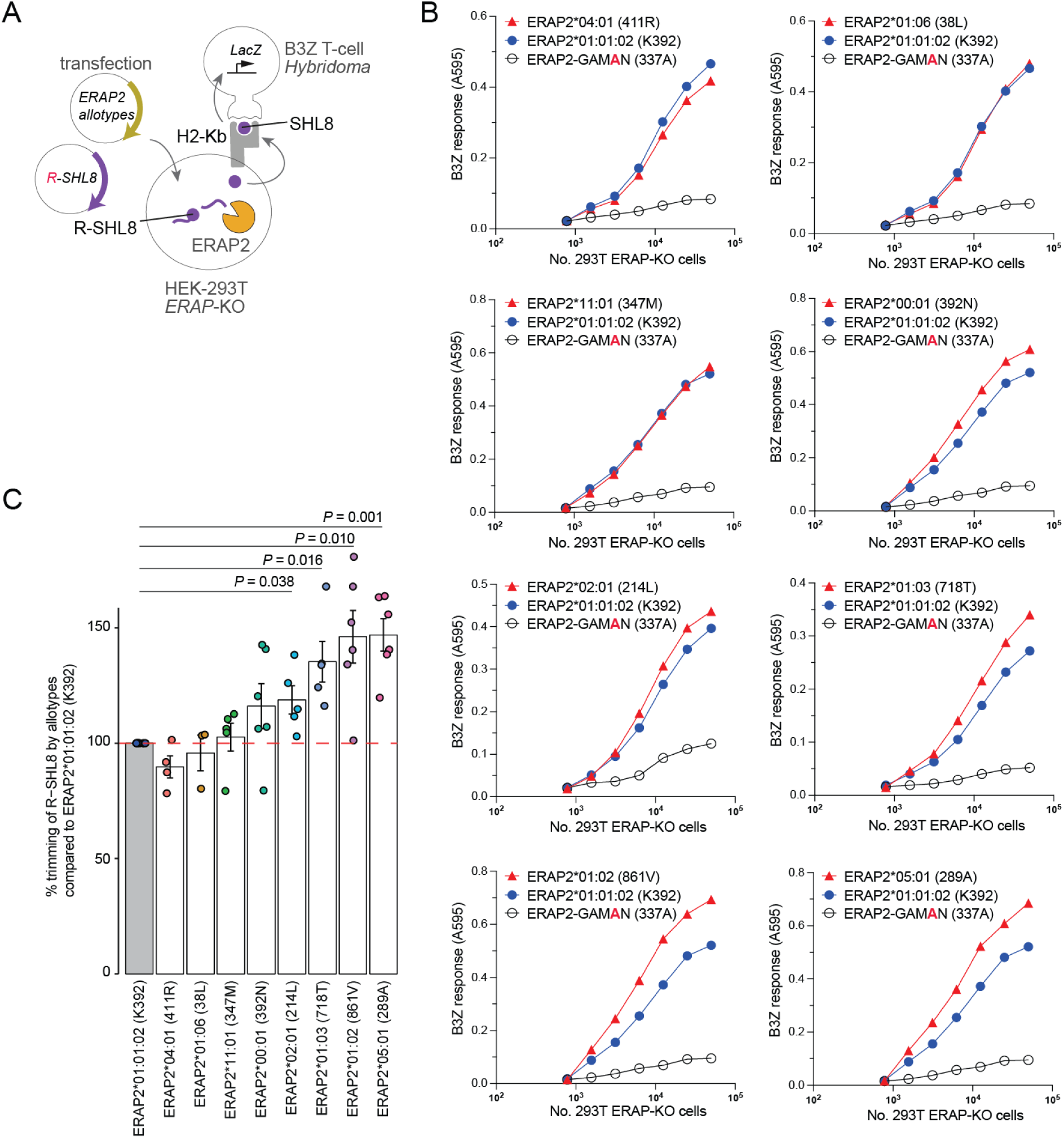
Disease-associated ERAP2 allotypes have more efficient peptide trimming activity. **A**) Schematic representation of the cell-based antigen presentation assay. ERAP-knockout HEK-293T cells were transfected with ERAP2 allotypes and extended precursor SIINFEHL (R-SHL8) and co-cultured with the H2-K^b^ restricted SHL8-specific, Lac-Z inducible T cell hybridoma cell line B3Z. Trimming of R-SHL8 by ERAP2 allotypes was indirectly measured through B3Z activation and the induced CPRG colour change. **B**) B3Z response to different effector:target ratios of HEK-293T cells transfected with selected ERAP2 allotypes, or the major ERAP2 allotype *ERAP2*01:01:02*, or an ERAP2 allotype without aminopeptidase activity (ERAP2-GAMAN). Shown is one representative of 3-6 independent repeats for tested ERAP2 allotype. **C**) Overall percentage trimming activity of ERAP2 allotypes compared to the major ERAP2 allotype *ERAP2*01:01:02* as 100%. Data is pooled from 3-6 independent repeats +/-SEM; P values are from a one-sample t-test.

## Discussion

This study demonstrates that the *ERAP2* gene is polymorphic in the European population and segregates into three major haplotype groups A, B, and C. These haplotypes give rise to functionally distinct ERAP2 protein allotypes that are associated with specific diseases in the population. The results of our study support the hypothesis that the biological functions of ERAP2 should be attributed to multiple allotypes, and that the widely believed dichotomous haplotype model of ERAP2 may not accurately capture the full extent of ERAP2 biology.

By using reference-free phasing, we aimed to map the *ERAP2* haplotype structure in the European population. While this approach may be more sensitive to detecting novel haplotypes since it is not biased by reference haplotypes, phasing based on low-frequency variants has an inherent trade-off in accuracy. Our mitigation strategy involved phasing the haplotypes using variants with an allele frequency down to 0.1% and we used imputation for rarer variants. Since imputation accuracy may decrease for very rare variants, we conservatively restricted our analysis to (well-annotated) variants with an allele frequency limited to 1:10,000, and used a sufficiently large sample size to ensure accurate phasing (43). By using relatively conservative thresholds, we ascertain that the haplotypes associated with these variants are robust, but a consequence of this approach is that our study did not provide an exhaustive catalogue of all epidemiologically relevant *ERAP2* haplotypes. Additional haplotypes containing other key amino acid substitutions may contribute to other diseases and require haplotype-based analysis in well powered case-control studies and using haplotype construction methods designed for very rare variant phasing based on long-range LD modelling (e.g., *SHAPEIT5* (43)) together with whole-genome sequencing data from a more diverse set of reference panels representing multiple populations. We recently showed that the haplotype-phased data of the 1000 Genomes Project (n∼2500) contains additional ERAP2 protein allotypes that persist with altered frequencies across continental populations (1), offering substantial room for future research to gain a deeper understanding of ERAP2 allotype diversity and its implications for human immunity. These follow-up studies will benefit from the availability of data from sources such as the Japan Biobank, Qatar Biobank (Middle Eastern ancestry), African Biobank and The All of Us Research Program by the National Institutes of Health. Such studies will benefit from complementary analysis using long-read mRNA-sequencing to map the full isoform usage of haplotypes of *ERAP2*. Another limitation is that we used the exome-sequencing data of the UK Biobank, and not the whole-genome sequencing data for phasing the haplotypes. This resulted in the manual assignment of the splice variant rs17486481 to haplotypes containing the variant 669Q, because the deep intronic variant rs17486481 was not covered in the exome-sequencing data. Another limitation of our study is that we assumed a significant proportion of the haplotypes do not produce (full-length) protein, because of the presence of splice-altering variants. Yet, previous studies show that a shorter ERAP2 transcript may be produced by ‘loss-of-function’ haplotypes (with the G allele of rs2248374) under specific inflammatory conditions (1, 15–17, 44). It would be interesting to determine if any or all of the here-reported haplotypes that are expected to be subjected to NMD also drive the expression of truncated transcripts or proteins during inflammation.

A key regulator of *ERAP2* expression is the SNV rs2248374 located inside the donor splice site of exon 10. The variant disrupts the canonical splice motif (13) and extends exon 10 to a downstream cryptic splice site that forces premature termination codons into the transcript, which effectively promotes nonsense-mediated mRNA degradation and causes loss of ERAP2 expression in haplotypes containing the G allele of rs2248374 (10). A plausible theory for the evolutionary selection for the G allele of rs2248374 is that it is in LD with the T allele of rs2549782 that encodes an asparagine at position 392 (392N), which significantly alters the substrate specificity of ERAP2 and results in a hyper-active enzyme for many substrates (39, 40). Note that our T cell activation assay did not detect differences between the ancestral allotype and the major ERAP2 allotype in the contemporary population, indicating that the 392N does not affect proteolytic activity towards the particular substrates used in this assay. Furthermore, our data suggests that by alternative splicing of rs2248374, also other protein-altering variants are precluded from expression, including haplotypes defined by other amino acid residues, including the alleles belonging to the *ERAP2*06*, *ERAP2*08, ERAP2*12*. Our data also supports selection against the A allele of missense variant rs17408150 (669Q), which we show significantly alters the enzymatic activity of ERAP2. While we have no functional data to support that this second splice variant functions similar to rs2248374 and leads to NMD, the variant is strongly associated with decreased ERAP2 expression independently from rs2248374 (10, 45), which suggests that it functions similarly by promoting NMD.

A variety of severe human diseases have been linked to ERAP2 gene variants, including several chronic inflammatory and autoimmune conditions (1, 8–10). Lethal infectious diseases (4, 7, 14), and cardiovascular complications (11, 12, 46–48). Additionally, novel treatments linked to altered outcomes of cancer immunotherapy are implicating ERAP2 in an increasing number of conditions (3, 49). Although this study used limited phenotype data from the UK Biobank, we demonstrated proof of concept that chronic inflammation, infectious disease and cardiovascular disease showed allotype-specific disease associations and strongly suggest that prior disease associations with ERAP2 with common variants stems from allotype-driven associations in diseases and the functional diversity of ERAP2. It will be necessary to functionally model allotypes in relevant cell-based assays or animal models in order to dissect the mechanisms of action.

The results of our study support the hypothesis that the biological functions of ERAP2 should be attributed to distinct protein-coding haplotypes, or allotypes, and that the widely believed dichotomous haplotype model of ERAP2 may not accurately capture the role of ERAP2 in human biology. Furthermore, the diversity in *ERAP2* haplotypes mimic similar findings in the closely related *ERAP1* gene. It has been shown that missense SNVs in *ERAP1* form multiple haplotypes in humans with varying enzymatic activities toward peptide substrates and differentially shape the immunopeptidome of MHC-I (9, 21–23). A number of these ERAP1 allotypes show disease-specific associations (9, 20, 23, 24, 50, 51). This suggests that both ERAP1 and ERAP2 are functionally distinct allotypes that broaden human immune variation.

In line with previous studies, we found an association between *ERAP2* and autoimmune conditions such as psoriatic arthritis, systemic sclerosis, and ankylosing spondylitis (52–55). Additionally, we found novel associations between specific *ERAP2* haplotypes and disorders of the cardiovascular system, venous thromboembolism, and secondary pulmonary hypertension which indicates that *ERAP2* may also function beyond antigen processing for MHC-I. ERAP2 has been reported to cleave angiotensin (ANG) II into ANG III and IV, suggesting its involvement in RAS pathways upon secretion in an extracellular environment (56). SNV in *ERAP2* have also been associated with blood pressure in previous GWAS studies (47, 48). In addition to these findings, we also found novel associations of *ERAP2* haplotypes with neurological disorders like migraine (*ERAP2*01:02*) and epilepsy (*ERAP2*05:01*). While neuro-inflammatory pathways are assumed to be implicated in the pathophysiology of both migraine and epilepsy, it remains currently unknown how ERAP2 allotypes contribute to these conditions (63). The here-identified allotypes may have important implications for other human pathologies. For example, ERAP2 has been linked to treatment efficacy and prognosis in cancer studies (14). Indeed, chemical inhibition of ERAP2 in a cancer cell line has been shown to shift the immunopeptidome to include many novel and potentially immunogenic peptides, suggesting potential applications in cancer immunotherapy (60). The MHC-I pathway plays a crucial role in vaccine efficacy and transplantation, so distinct ERAP2 allotypes may also play a role in these areas.

In conclusion, our study highlights the substantial protein diversity of ERAP2 within the human population, showing significant functional implications for disease predisposition. These findings provide valuable insight into disease mechanisms in which ERAP2 activity may be a viable target for drug development, particularly in autoimmune, and other inflammatory disorders.

## Data Availability

All data produced are available online at https://github.com/Araja1234/ERAP2-Haplotypes-Analysis-UK-Biobank/

## Acknowledgements

This project has received funding from the European Union’s Horizon 2020 research and innovation programme under the Marie Skłodowska-Curie grant agreement No 954992. <u>This research has been</u> <u>conducted using the UK Biobank Resource under Application Number 24711.</u>

## Supplementary Figures

**Supplementary Figure 1.**
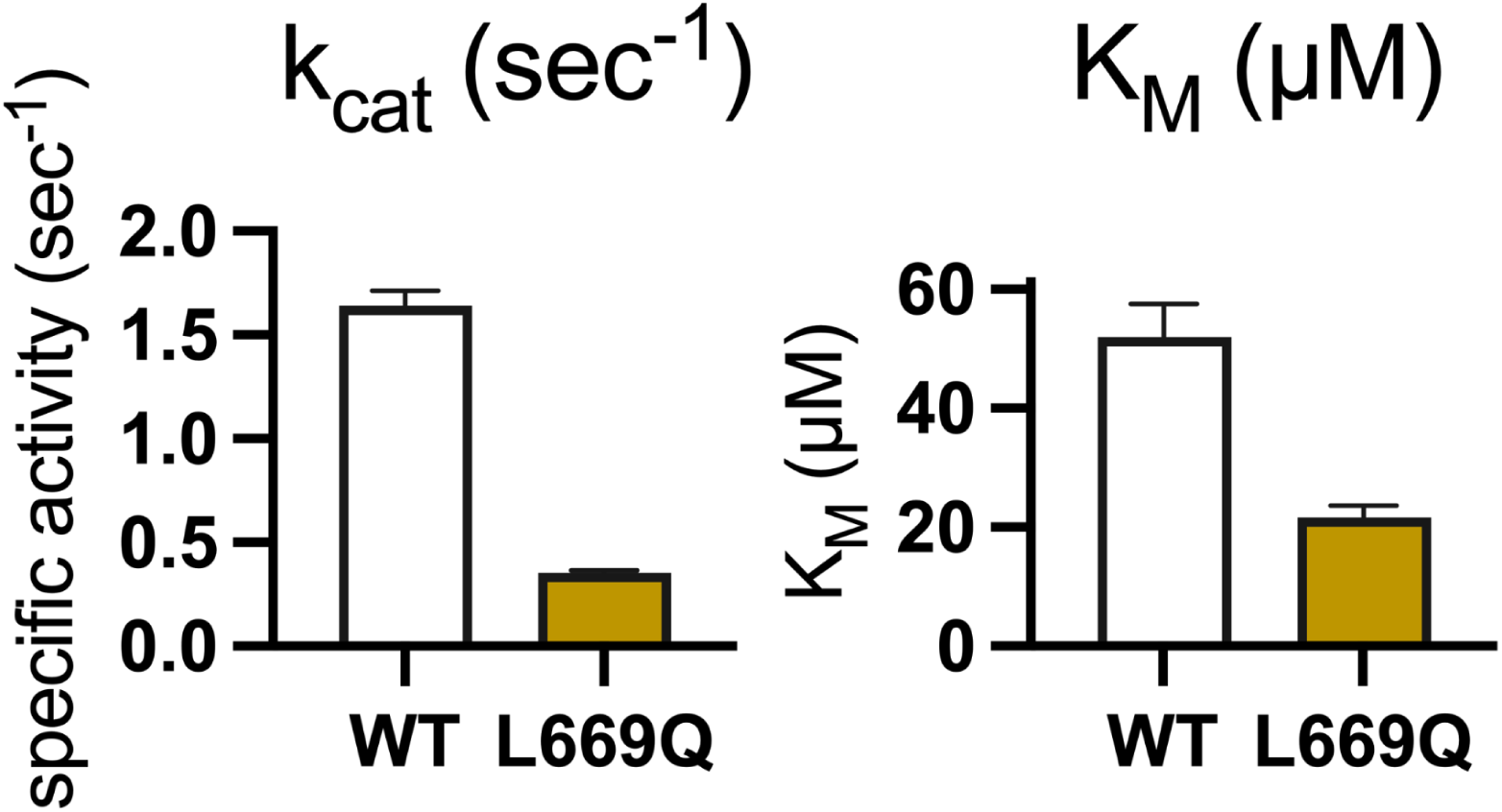
Supplemental Figure to Figure 1F. **Left Panel.** The calculated catalytic turnover for ERAP2 WT (669L) and the amino acid substitution L669Q (669Q). **Right Panel.** The calculated K_M_ constant for ERAP2 WT (669L) and the amino acid substitution L669Q (669Q).

**Supplementary Figure 2:**
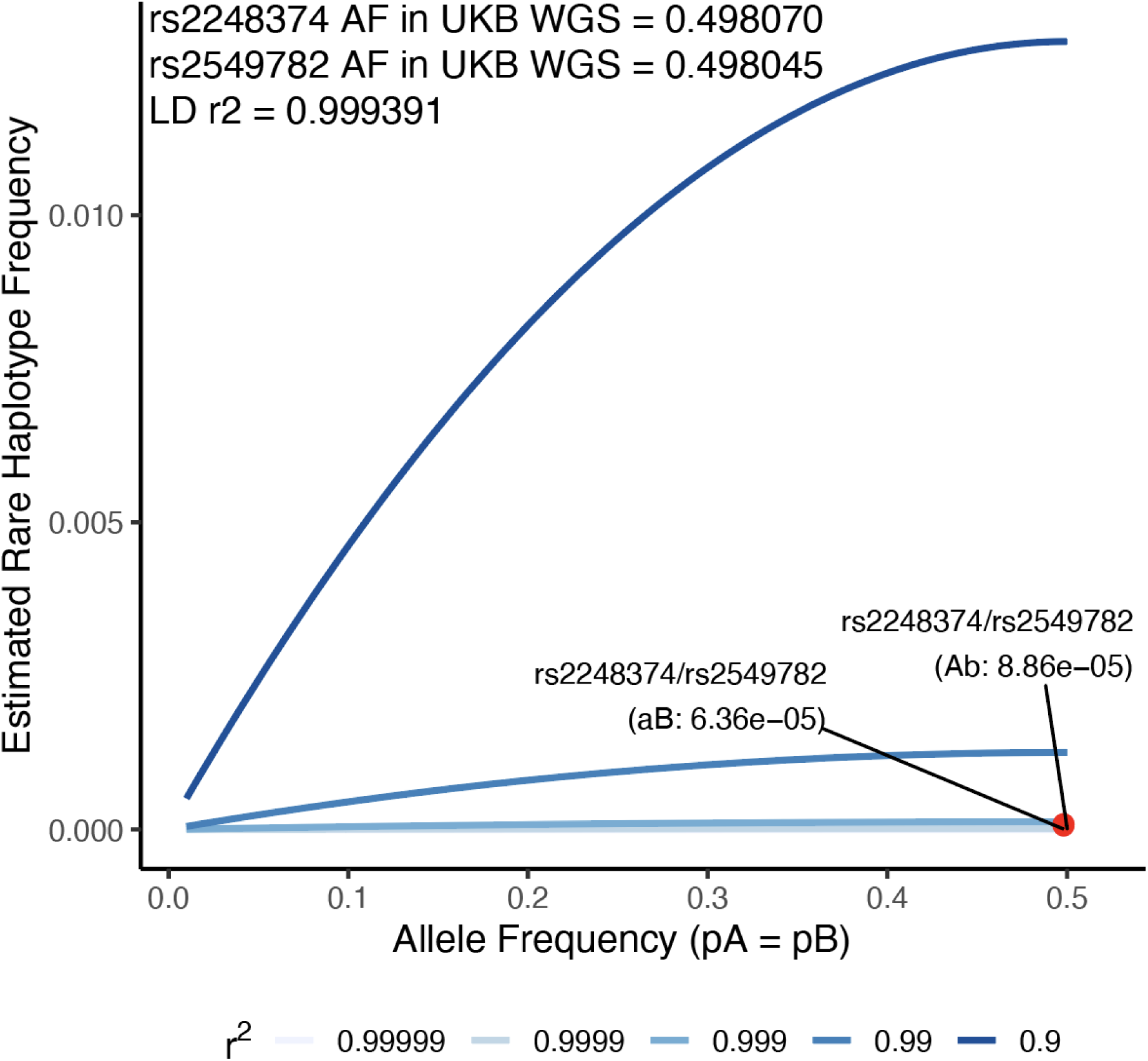
Estimation of the ancestral haplotype frequency of rs2248374 and rs2549782 in the European Population. The expected frequency of rare haplotypes under varying levels of LD (r^2^) and allele frequencies were simulated. The specific points highlighted correspond to the estimated frequency of the rare ancestral haplotype *ERAP2*00:01* of rs2248374-rs2549782 (Ab) and *ERAP2*01:01:01* (aB) based on allele frequencies and LD (r^2^) between these variants in the UK Biobank WGS data.

**Supplementary Table 1.** Association of *ERAP2* haplotypes with 382 ICD-10 (International Classification of Disease, 10th Revision) codes in the UK Biobank. The association of *ERAP2* haplotypes with the phenotypes was assessed by logistic regression model and *P* values corrected using the Benjamini–Hochberg (BH) method.

| *ERAP2*<br>Haplotype | Frequency |  | ICD-10 condition<br>(code) | Adjusted P<br>value | Odds ratio [95%<br>confidence<br>interval |
| --- | --- | --- | --- | --- | --- |
|  | Case | Control |  |  |  |
| *ERAP2* *01:01:02 | 0.427 | 0.439 | venous<br>thromboembolism | $1.87 \times 10^{-2}$ | 0.951 (0.918, 0.986) |
| *ERAP2* *01:01:02 | 0.377 | 0.439 | Dilated<br>cardiomyopathy<br>(PH145) | $5.07 \times 10^{-3}$ | 0.767 (0.650, 0.905) |
| *ERAP2* *01:01:02 | 0.533 | 0.439 | Systemic sclerosis<br>(PH318) | $3.24 \times 10^{-2}$ | 1.461 (1.092, 1.955) |
| *ERAP2* *01:01:02 | 0.507 | 0.439 | Trigeminal neuralgia<br>(PH325) | $1.37 \times 10^{-2}$ | 1.318 (1.089, 1.596) |
| *ERAP2* *01:01:02 | 0.405 | 0.439 | Multiple sclerosis<br>(PH63) | $4.44 \times 10^{-2}$ | 0.866 (0.772, 0.972) |
| *ERAP2* *01:01:02 | 0.490 | 0.439 | Psoriatic arthropathy<br>(PH75) | $1.41 \times 10^{-2}$ | 1.233 (1.066, 1.427) |
| *ERAP2* *01:02 | 0.003 | $9.7 \times 10^{-4}$ | Migraine (PH211) | $9.42 \times 10^{-3}$ | 2.885 (1.428, 5.828) |
| *ERAP2* *01:03 | 0.001 | $7.7 \times 10^{-4}$ | venous<br>thromboembolism | $2.02 \times 10^{-2}$ | 1.906 (1.195, 3.040) |
| *ERAP2* *02:01 | $3.7 \times 10^{-4}$ | 0.001 | Infection of skin and<br>subcutaneous tissues<br>(PH308) | $2.55 \times 10^{-2}$ | 0.257 (0.094, 0.707) |
| *ERAP2* *05:01 | 0.003 | 0.001 | Nonalcoholic fatty liver<br>disease | $1.11 \times 10^{-2}$ | 3.213 (1.461, 7.064) |
| *ERAP2* *05:01 | 0.003 | 0.001 | Endometrial<br>hyperplasia and<br>hypertrophy (PH161) | $2.32 \times 10^{-2}$ | 2.945 (1.330, 6.521) |
| *ERAP2* *05:01 | 0.003 | 0.001 | Epilepsy (PH165) | $1.42 \times 10^{-2}$ | 2.536 (1.330, 4.839) |
| *ERAP2* *05:01 | 0.004 | 0.001 | Fatty Liver (PH168) | $5.20 \times 10^{-3}$ | 3.252 (1.555, 6.801) |
| *ERAP2* *05:01 | 0.004 | 0.001 | Primary Malignancy<br>Uterine (PH271) | $1.20 \times 10^{-2}$ | 3.697 (1.518, 9.004) |
| *ERAP2* *06:01 | | | venous<br>thromboembolism | $4.15 \times 10^{-3}$ | 1.059 (1.023, 1.097) |
|  | 0.473 | 0.459 |  |  |  |
| ERAP2*06:01 | 0.474 | 0.459 | Pulmonary embolism<br>(PH978 / 2156) | $4.40 \times 10^{-2}$ | 1.064 (1.012, 1.119) |
| ERAP2*06:01 | 0.363 | 0.459 | Systemic sclerosis<br>(PH318) | $2.52 \times 10^{-2}$ | 0.666 (0.492, 0.901) |
| ERAP2*06:01 | 0.398 | 0.459 | Trigeminal neuralgia<br>(PH325) | $3.52 \times 10^{-2}$ | 0.778 (0.640, 0.946) |
| ERAP2*06:01 | 0.482 | 0.459 | Venous_thromboemb<br>olic (PH338) | $2.33 \times 10^{-2}$ | 1.096 (1.024, 1.172) |
| ERAP2*06:01 | 0.415 | 0.459 | Psoriatic arthropathy<br>(PH75) | $4.55 \times 10^{-2}$ | 0.833 (0.719, 0.965) |
| ERAP2*08:01 | 0.023 | 0.034 | Nonrheumatic aortic<br>valve disorders<br>(PH219) | $3.18 \times 10^{-3}$ | 0.653 (0.506, 0.843) |
| ERAP2*08:01 | 0.055 | 0.034 | Ankylosing spondylitis<br>(PH99) | $1.95 \times 10^{-2}$ | 1.664 (1.153, 2.403) |
| ERAP2*10:01 | 0.081 | 0.056 | Secondary pulmonary<br>hypertension(PH300) | $4.23 \times 10^{-2}$ | 1.491 (1.084, 2.051) |

**Supplementary Table 2:**
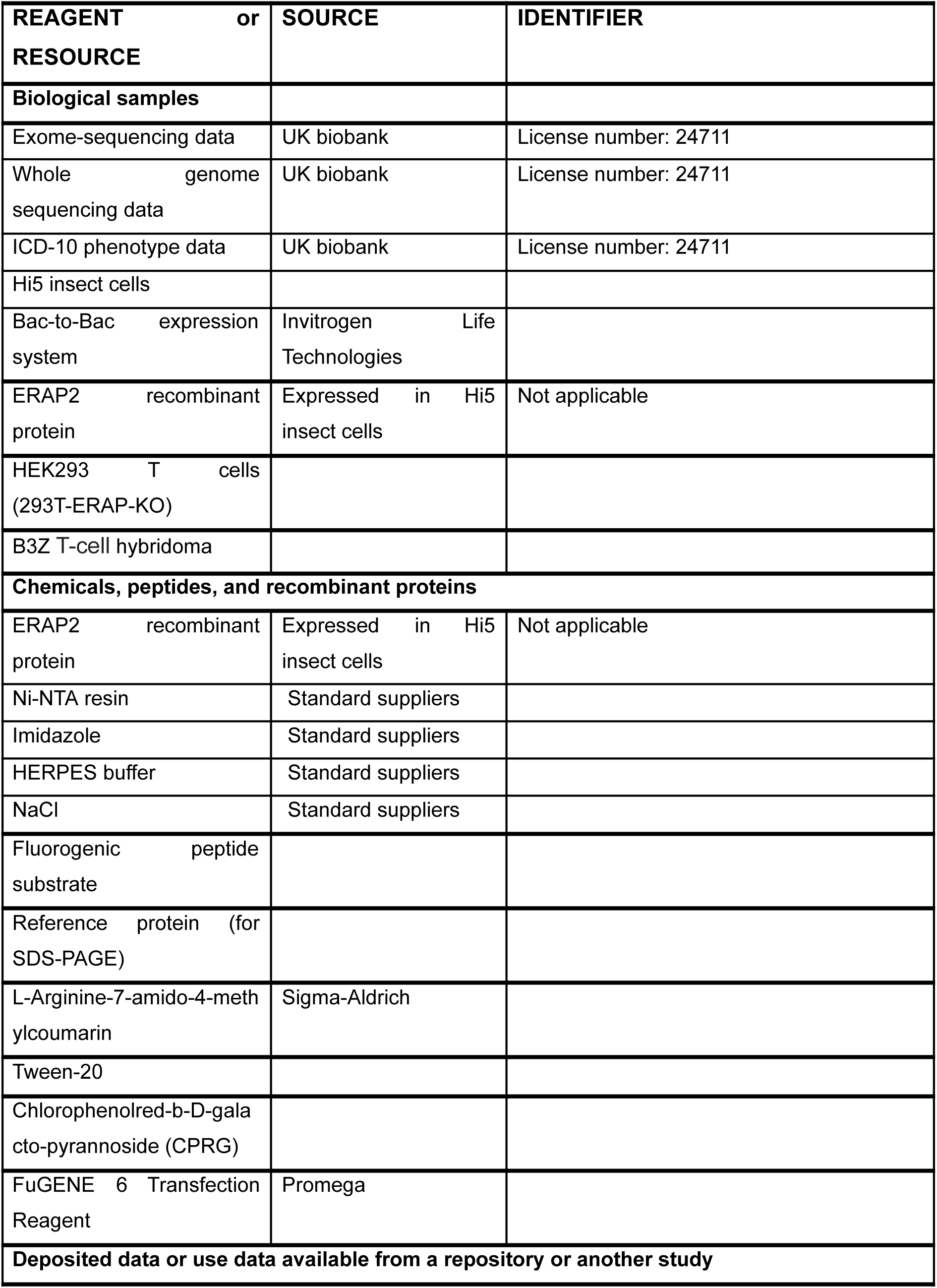
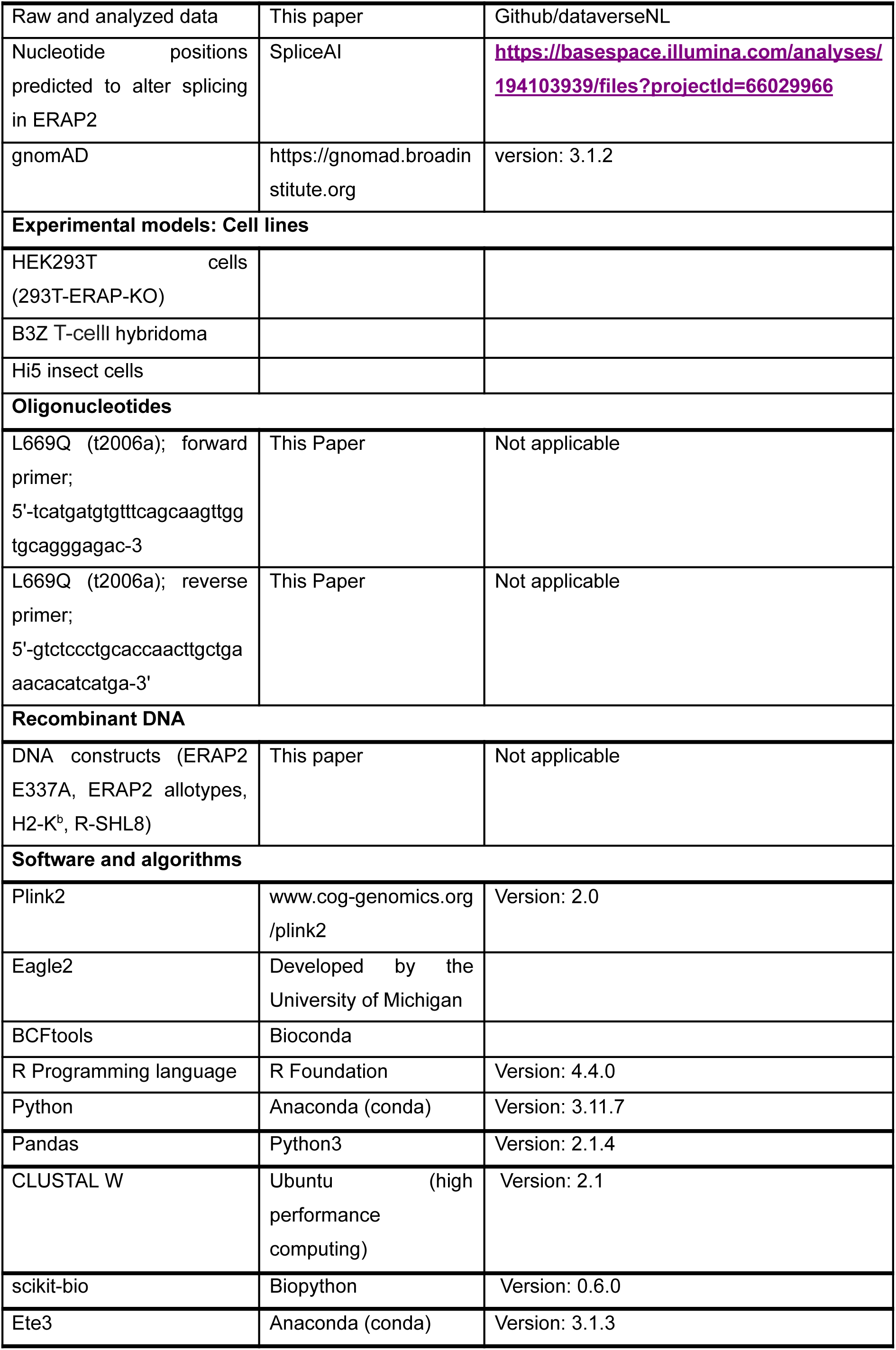
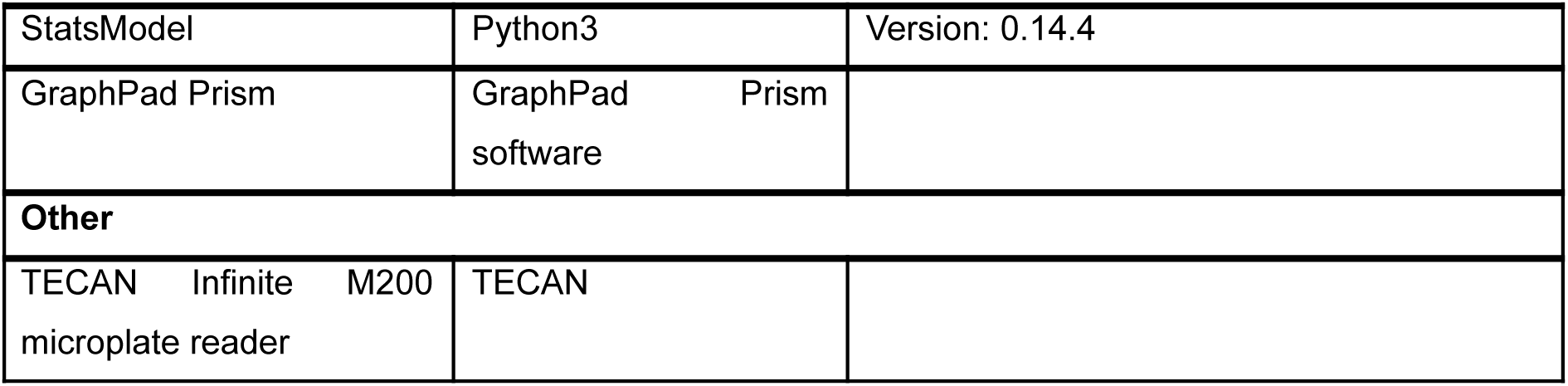
Key resource Table including details of all the reagents, biological samples, data, chemicals, peptides, recombinant proteins, software, Python and R packages used in this study alongside their sources and identifiers where applicable.

**Supplementary Table 3:** *ERAP2* haplotypes count and frequency in UK Biobank data.

| Haplotype | Count | Raw Frequency | Frequency (%) |
| --- | --- | --- | --- |
| ERAP2*06:01 | 153428 | 0.46 | 46 |
| ERAP2*01:01:02 | 146582 | 0.44 | 44 |
| ERAP2*10:01 | 18814 | 0.056 | 5.6 |
| ERAP2*08:01 | 11356 | 0.034 | 3.4 |
| ERAP2*04:01 | 998 | 0.003 | 0.3 |
| ERAP2*02:01 | 445 | 0.0013 | 0.13 |
| ERAP2*05:01 | 376 | 0.0011 | 0.11 |
| ERAP2*06:02 | 367 | 0.00109 | 0.109 |
| ERAP2*06:03 | 361 | 0.00108 | 0.108 |
| ERAP2*01:02 | 329 | 0.00099 | 0.099 |
| ERAP2*01:03 | 267 | 0.000799 | 0.0799 |
| ERAP2*06:04 | 251 | 0.000752 | 0.0752 |
| ERAP2*06:05 | 103 | 0.000308 | 0.0308 |
| ERAP2*06:06 | 100 | 0.000299 | 0.0299 |
| ERAP2*01:04 | 67 | 0.0002006 | 0.02006 |
| ERAP2*10:02 | 51 | 0.0001527 | 0.01527 |
| ERAP2*01:01:01 | 31 | $9.28 \times 10^{-5}$ | 0.00928 |
| ERAP2*11:01 | 28 | $8.384 \times 10^{-5}$ | 0.008384 |
| ERAP2*00:01 | 8 | $2.395 \times 10^{-5}$ | 0.002395 |
| ERAP2*06:07 | 5 | $1.4971 \times 10^{-5}$ | 0.0014971 |
| ERAP2*11:02 | 4 | $1.1977 \times 10^{-5}$ | 0.0011977 |
| ERAP2*01:06 | 3 | $8.983 \times 10^{-6}$ | 0.0008983 |
| *ERAP2\*06:08* | 2 | 5.9884 10<sup>-6</sup> | 0.00059884 |
| *ERAP2\*08:02* | 1 | 2.9942 10<sup>-6</sup> | 0.00029942 |
| *ERAP2\*12:01* | 1 | 2.9942 10<sup>-6</sup> | 0.00029942 |
| *ERAP2\*06:09* | 1 | 2.9942 10<sup>-6</sup> | 0.00029942 |
| *ERAP2\*10:03* | 1 | 2.9942 10<sup>-6</sup> | 0.00029942 |

## References

1. A. Raja, J. J. W. Kuiper, Evolutionary immuno-genetics of endoplasmic reticulum aminopeptidase II (ERAP2). Genes Immun. 24, 295–302 (2023).

2. W. J. Venema, et al., ERAP2 Increases the Abundance of a Peptide Submotif Highly Selective for the Birdshot Uveitis-Associated HLA-A29. Front. Immunol. 12, 634441 (2021).

3. H.-W. Tsao, et al., Targeting the aminopeptidase ERAP enhances antitumor immunity by disrupting the NKG2A-HLA-E inhibitory checkpoint. Immunity 57, 2863–2878.e12 (2024).

4. J. Klunk, et al., Evolution of immune genes is associated with the Black Death. Nature 611, 312–319 (2022).

5. G. Kerner, et al., Genetic adaptation to pathogens and increased risk of inflammatory disorders in post-Neolithic Europe. Cell Genomics 3, 100248 (2023).

6. G. Laval, E. Patin, L. Quintana-Murci, G. Kerner, Deep estimation of the intensity and timing of natural selection from ancient genomes. Mol. Ecol. Resour. 24, e14015 (2024).

7. R. Cagliani, et al., Genetic diversity at endoplasmic reticulum aminopeptidases is maintained by balancing selection and is associated with natural resistance to HIV-1 infection. Hum. Mol. Genet. 19, 4705–4714 (2010).

8. F. Hamilton, et al., Variation in ERAP2 has opposing effects on severe respiratory infection and autoimmune disease. Am. J. Hum. Genet. 110, 691–702 (2023).

9. J. J. Kuiper, et al., EULAR study group on “MHC-I-opathy”: identifying disease-overarching mechanisms across disciplines and borders. Ann. Rheum. Dis. 82, 887–896 (2023).

10. W. J. Venema, et al., A cis-regulatory element regulates ERAP2 expression through autoimmune disease risk SNPs. Cell Genomics 4, 100460 (2024).

11. L. D. Hill, et al., Fetal ERAP2 variation is associated with preeclampsia in African Americans in a case-control study. BMC Med. Genet. 12, 64 (2011).

12. J. M. Rimpelä, et al., Genome-wide association study of nocturnal blood pressure dipping in hypertensive patients. BMC Med. Genet. 19, 110 (2018).

13. A. M. Andrés, et al., Balancing selection maintains a form of ERAP2 that undergoes nonsense-mediated decay and affects antigen presentation. PLoS Genet. 6, e1001157 (2010).

14. A. L. Hanson, et al., Genetic Variants in ERAP1 and ERAP2 Associated With Immune-Mediated Diseases Influence Protein Expression and the Isoform Profile. Arthritis Rheumatol. Hoboken NJ 70, 255–265 (2018).

15. C. J. Ye, et al., Genetic analysis of isoform usage in the human anti-viral response reveals influenza-specific regulation of ERAP2 transcripts under balancing selection. Genome Res. 28, 1812–1825 (2018).

16. I. Saulle, et al., A New ERAP2/Iso3 Isoform Expression Is Triggered by Different Microbial Stimuli in Human Cells. Could It Play a Role in the Modulation of SARS-CoV-2 Infection? Cells 9, 1951 (2020).

17. B. Mattorre, et al., A Short ERAP2 That Binds IRAP Is Expressed in Macrophages Independently of Gene Variation. Int. J. Mol. Sci. 23, 4961 (2022).

18. K. Hamazaki, H. Iwata, RAINBOW: Haplotype-based genome-wide association study using a novel SNP-set method. PLoS Comput. Biol. 16, e1007663 (2020).

19. S. P. Dickson, K. Wang, I. Krantz, H. Hakonarson, D. B. Goldstein, Rare variants create synthetic genome-wide associations. PLoS Biol. 8, e1000294 (2010).

20. J. P. Hutchinson, et al., Common allotypes of ER aminopeptidase 1 have substrate-dependent and highly variable enzymatic properties. J. Biol. Chem. 296, 100443 (2021).

21. M. J. Ombrello, D. L. Kastner, E. F. Remmers, Endoplasmic reticulum-associated amino-peptidase 1 and rheumatic disease: genetics. Curr. Opin. Rheumatol. 27, 349–356 (2015).

22. J. A. López de Castro, How ERAP1 and ERAP2 Shape the Peptidomes of Disease-Associated MHC-I Proteins. Front. Immunol. 9, 2463 (2018).

23. A. Arakawa, et al., ERAP1 Controls the Autoimmune Response against Melanocytes in Psoriasis by Generating the Melanocyte Autoantigen and Regulating Its Amount for HLA-C*06:02 Presentation. J. Immunol. Baltim. Md 1950 207, 2235–2244 (2021).

24. S. Gelfman, et al., A large meta-analysis identifies genes associated with anterior uveitis. Nat. Commun. 14, 7300 (2023).

25. J. J. W. Kuiper, et al., Functionally distinct ERAP1 and ERAP2 are a hallmark of HLA-A29-(Birdshot) Uveitis. Hum. Mol. Genet. 27, 4333–4343 (2018).

26. A. R. Roberts, et al., ERAP1 association with ankylosing spondylitis is attributable to common genotypes rather than rare haplotype combinations. Proc. Natl. Acad. Sci. U. S. A. 114, 558–561 (2017).

27. C. V. Van Hout, et al., Exome sequencing and characterization of 49,960 individuals in the UK Biobank. Nature 586, 749–756 (2020).

28. S. Chen, et al., A genomic mutational constraint map using variation in 76,156 human genomes. Nature 625, 92–100 (2024).

29. S. Purcell, et al., PLINK: a tool set for whole-genome association and population-based linkage analyses. Am. J. Hum. Genet. 81, 559–575 (2007).

30. J. Cheng, et al., Accurate proteome-wide missense variant effect prediction with AlphaMissense. Science 381, eadg7492 (2023).

31. M. J. Machiela, S. J. Chanock, LDlink: a web-based application for exploring population-specific haplotype structure and linking correlated alleles of possible functional variants. Bioinforma. Oxf. Engl. 31, 3555–3557 (2015).

32. J. D. Thompson, D. G. Higgins, T. J. Gibson, CLUSTAL W: improving the sensitivity of progressive multiple sequence alignment through sequence weighting, position-specific gap penalties and weight matrix choice. Nucleic Acids Res. 22, 4673–4680 (1994).

33. N. Saitou, M. Nei, The neighbor-joining method: a new method for reconstructing phylogenetic trees. Mol. Biol. Evol. 4, 406–425 (1987).

34. J. Huerta-Cepas, F. Serra, P. Bork, ETE 3: Reconstruction, Analysis, and Visualization of Phylogenomic Data. Mol. Biol. Evol. 33, 1635–1638 (2016).

35. S. Seabold, J. Perktold, statsmodels: Econometric and statistical modeling with python in 9th Python in Science Conference, (2010).

36. J. R. Birtley, E. Saridakis, E. Stratikos, I. M. Mavridis, The crystal structure of human endoplasmic reticulum aminopeptidase 2 reveals the atomic basis for distinct roles in antigen processing. Biochemistry 51, 286–295 (2012).

37. E. Reeves, C. J. Edwards, T. Elliott, E. James, Naturally occurring ERAP1 haplotypes encode functionally distinct alleles with fine substrate specificity. J. Immunol. Baltim. Md 1950 191, 35–43 (2013).

38. C. M. Brand, L. L. Colbran, J. A. Capra, Resurrecting the alternative splicing landscape of archaic hominins using machine learning. *Nat*. Ecol. Evol. 7, 939–953 (2023).

39. I. Evnouchidou, et al., A common single nucleotide polymorphism in endoplasmic reticulum aminopeptidase 2 induces a specificity switch that leads to altered antigen processing. J. Immunol. Baltim. Md 1950 189, 2383–2392 (2012).

40. D. Forni, et al., An evolutionary analysis of antigen processing and presentation across different timescales reveals pervasive selection. PLoS Genet. 10, e1004189 (2014).

41. A. I. Marusina, et al., Cell-Specific and Variant-Linked Alterations in Expression of ERAP1, ERAP2, and LNPEP Aminopeptidases in Psoriasis. J. Invest. Dermatol. 143, 1157–1167.e10 (2023).

42. J. Ghouse, et al., Genome-wide meta-analysis identifies 93 risk loci and enables risk prediction equivalent to monogenic forms of venous thromboembolism. Nat. Genet. 55, 399–409 (2023).

43. R. J. Hofmeister, D. M. Ribeiro, S. Rubinacci, O. Delaneau, Accurate rare variant phasing of whole-genome and whole-exome sequencing data in the UK Biobank. Nat. Genet. 55, 1243–1249 (2023).

44. Y. Zheng, N. Goulter, Introduction to the Special Issue: Novel Insights into the Externalizing Psychopathology Spectrum in Childhood and Adolescence from Intensive Longitudinal Data. Res. Child Adolesc. Psychopathol. 52, 1–6 (2024).

45. B. B. Sun, et al., Genomic atlas of the human plasma proteome. Nature 558, 73–79 (2018).

46. M. P. Johnson, et al., The ERAP2 gene is associated with preeclampsia in Australian and Norwegian populations. Hum. Genet. 126, 655–666 (2009).

47. S. Sakaue, et al., A cross-population atlas of genetic associations for 220 human phenotypes. Nat. Genet. 53, 1415–1424 (2021).

48. P. Surendran, et al., Discovery of rare variants associated with blood pressure regulation through meta-analysis of 1.3 million individuals. Nat. Genet. 52, 1314–1332 (2020).

49. Y. W. Lim, et al., Germline genetic polymorphisms influence tumor gene expression and immune cell infiltration. Proc. Natl. Acad. Sci. U. S. A. 115, E11701–E11710 (2018).

50. M. Takeuchi, et al., A single endoplasmic reticulum aminopeptidase-1 protein allotype is a strong risk factor for Behçet’s disease in HLA-B*51 carriers. Ann. Rheum. Dis. 75, 2208–2211 (2016).

51. T. Rayinda, et al., Epistasis of ERAP1 With 4 Major Histocompatibility Complex Class I Alleles in Frontal Fibrosing Alopecia: A Genome-Wide Association Study Meta-Analysis. JAMA Dermatol. 161, 310–314 (2025).

52. M. Kerick, et al., Expression Quantitative Trait Locus Analysis in Systemic Sclerosis Identifies New Candidate Genes Associated With Multiple Aspects of Disease Pathology. Arthritis Rheumatol. Hoboken NJ 73, 1288–1300 (2021).

53. P. C. Robinson, et al., ERAP2 is associated with ankylosing spondylitis in HLA-B27-positive and HLA-B27-negative patients. Ann. Rheum. Dis. 74, 1627–1629 (2015).

54. A. Fierabracci, A. Milillo, F. Locatelli, D. Fruci, The putative role of endoplasmic reticulum aminopeptidases in autoimmunity: insights from genomic-wide association studies. Autoimmun. Rev. 12, 281–288 (2012).

55. Y. Luo, et al., Cross-Phenotype GWAS Supports Shared Genetic Susceptibility to Systemic Sclerosis and Primary Biliary Cholangitis. MedRxiv Prepr. Serv. Health Sci. 2024.07.01.24309721 (2024). 10.1101/2024.07.01.24309721.

56. B. Mattorre, et al., The emerging multifunctional roles of ERAP1, ERAP2 and IRAP between antigen processing and renin-angiotensin system modulation. Front. Immunol. 13, 1002375 (2022).

57. O. Kursun, M. Yemisci, A. M. J. M. van den Maagdenberg, H. Karatas, Migraine and neuroinflammation: the inflammasome perspective. J. Headache Pain 22, 55 (2021).

58. A. Rana, A. E. Musto, The role of inflammation in the development of epilepsy. J. Neuroinflammation 15, 144 (2018).

59. D. Fruci, et al., Altered expression of endoplasmic reticulum aminopeptidases ERAP1 and ERAP2 in transformed non-lymphoid human tissues. J. Cell. Physiol. 216, 742–749 (2008).

60. I. Temponeras, et al., ERAP2 Inhibition Induces Cell-Surface Presentation by MOLT-4 Leukemia Cancer Cells of Many Novel and Potentially Antigenic Peptides. Int. J. Mol. Sci. 23, 1913 (2022).

